# CLINICAL OPPORTUNITIES FOR GERMLINE PHARMACOGENETICS AND MANAGEMENT OF DRUG-DRUG INTERACTIONS IN PATIENTS WITH ADVANCED SOLID CANCERS

**DOI:** 10.1101/2021.08.23.21262496

**Authors:** Tyler Shugg, Reynold C. Ly, Elizabeth J. Rowe, Santosh Philips, Mustafa A. Hyder, Milan Radovich, Marc B. Rosenman, Victoria M. Pratt, John T. Callaghan, Zeruesenay Desta, Bryan P. Schneider, Todd C. Skaar

## Abstract

**PURPOSE:** Precision medicine approaches, including germline pharmacogenetics (PGx) and management of drug-drug interactions (DDIs), are likely to benefit advanced cancer patients who are frequently prescribed multiple concomitant medications to treat cancer and associated conditions. Our objective was to assess the potential opportunities for PGx and DDI management within a cohort of adults with advanced cancer.

**PATIENTS AND METHODS:** Medication data were collected from the electronic health records (EHRs) for 481 subjects since their first cancer diagnosis. All subjects were genotyped for variants with clinically actionable recommendations in Clinical Pharmacogenetics Implementation Consortium (CPIC) guidelines for 13 pharmacogenes. DDIs were defined as concomitant prescription of strong inhibitors or inducers with sensitive substrates of the same drug-metabolizing enzyme and were assessed for six major cytochrome P450 (CYP) enzymes.

**RESULTS:** Approximately 60% of subjects were prescribed at least one medication with CPIC recommendations, and ∼14% of subjects had an instance for actionable PGx, defined as prescription of a drug in a subject with an actionable genotype. The overall subject-level prevalence of DDIs and serious DDIs were 50.3% and 34.8%, respectively. Serious DDIs were most common for CYP3A, CYP2D6, and CYP2C19, occurring in 24.9%, 16.8%, and 11.7% of subjects, respectively. When assessing PGx and DDIs together, ∼40% of subjects had at least one opportunity for a precision medicine-based intervention and ∼98% of subjects had an actionable phenotype for at least one CYP enzyme.

**CONCLUSION:** Our findings demonstrate numerous clinical opportunities for germline PGx and DDI management in adults with advanced cancer.

## INTRODUCTION

Pharmacogenetics (PGx) and management of drug-drug interactions (DDIs) are two aspects of precision medicine that have the potential to optimize medication therapy in oncology and other therapeutic disciplines. PGx-guided approaches have been shown to enhance drug efficacy and safety, including results from prospective clinical trials that have demonstrated the potential for PGx to improve drug safety.^1-3^ Accordingly, the U.S. Food and Drug Administration (FDA) currently includes PGx information within the labels for nearly 300 medications.^4^ Moreover, clinical practice guidelines that include PGx-guided recommendations have been published by the Clinical Pharmacogenetics Implementation Consortium (CPIC) and prominent discipline-specific professional organizations (e.g., the National Comprehensive Cancer Network) for over 100 medications.^5,6^ Similarly, DDIs are known to contribute to adverse drug events,^7,8^ and strategies to manage DDIs have been shown to improve patient outcomes.^9^ Given their important clinical implications, DDIs constitute a major consideration both during drug development and in clinical medicine, and recommendations to manage DDIs are therefore included both in FDA drug development guidance to industry^10^ and in numerous clinical practice guidelines.^11,12^

The clinical utility of precision medicine is expected to be especially high for patients with advanced cancer given that drug therapy is commonly used not only to treat cancer, but also to manage both cancer treatment-related adverse events (e.g., nausea and vomiting) and comorbid conditions associated with cancer (e.g., psychiatric conditions and pain syndromes). As a result, polypharmacy, typically defined as the concomitant use of 5 or more drugs, is exceedingly common in advanced cancer patients.^13^ Polypharmacy carries an increased risk for DDIs,^14^ and, predictably, multiple investigations have identified serious DDIs in advanced cancer that impact patient outcomes.^15^ PGx-guided approaches also offer the ability to optimize therapy for numerous anticancer medications based on somatic and germline genetic biomarkers. While molecular tumor boards have effectively harnessed somatic genome-guided treatment approaches to improve patient outcomes,^16^ germline PGx biomarkers can enhance medication safety with agents such as fluoropyrimidine and thiopurine chemotherapies.^17,18^ Additionally, PGx-guided approaches have been shown to enhance both efficacy and safety of selective serotonin reuptake inhibitors (SSRIs), tricyclic antidepressants (TCAs), and opioid analgesics that are often prescribed for comorbid conditions prevalent in cancer.^19-21^ Given these abundant PGx opportunities in cancer patients, it has been suggested that preemptive testing for PGx variants at first cancer diagnosis may be an effective clinical strategy to optimize patient outcomes.^22^ Furthermore, recent advancements in bioinformatics technology have enhanced the feasibility of PGx approaches in cancer through the creation of methods to extract PGx information from existing germline sequencing data generated during the clinical workflow of molecular tumor boards.^23,24^

Although past studies have characterized opportunities for DDI management and PGx-guided approaches in patients with advanced cancer, we are not aware of any work that has simultaneously investigated both approaches to provide a comprehensive assessment of the potential for precision medicine. Therefore, the objective of this study was to determine composite opportunities for precision medicine, incorporating both PGx-guided and DDI management strategies, within a cohort of adults with advanced solid cancers. By analyzing the potential for PGx-guided interventions since each subject’s respective date of first cancer diagnosis, we also directly investigate the potential clinical utility of preemptively obtaining PGx information when patients are first diagnosed with cancer.

## METHODS

### Subject Enrollment and Eligibility

This study was a retrospective electronic health record (EHR) review and prospective genotyping of eligible patients with solid cancers at Indiana University Health in Indianapolis, Indiana, USA. Subjects were eligible to participate in the study if they 1) had been seen in the Indiana University Health Precision Genomics clinic and enrolled in the accompanying Indiana University Total Cancer Care Protocol (part of the larger Oncology Research Information Exchange Network-wide Total Cancer Care initiative [https://www.oriencancer.org/]) and 2) agreed to submit a blood sample for genotyping. Subjects were enrolled into the study at clinic visits from February 2015 to February 2018. This research protocol, as well as the parent Total Cancer Care Protocol, were approved by Indiana University’s Institutional Review Board. All subjects provided written informed consent.

### Study Design and Data Collection

The purpose of this study was to investigate potential opportunities for precision medicine interventions, including PGx and management of DDIs, within a cohort of 481 adults seen at our institutional precision oncology clinic and associated solid tumor board. Demographic and clinical data, including medication prescriptions, were collected from the EHRs of all institutions participating in the Indiana Health Information Exchange, a statewide EHR data repository that includes 38 healthcare systems. Demographic data included age, sex, and race. Clinical data included first oncologic diagnosis and all inpatient and outpatient prescriptions. Genotyping for major pharmacogenes was performed at the College of American Pathologists-accredited Indiana University Pharmacogenomics Laboratory using a laboratory-developed assay based on the OpenArray^®^ Platform (ThermoFisher; Waltham, MA). The genes included on the genotyping platform, along with the number of variants tested for each gene, were as follows: *CYP2B6* (2), *CYP2C19* (6), *CYP2C9* (6), *CYP2D6* (11, including copy number targeting exon 9), *CYP3A4* (2), *CYP3A5* (3), *CYP4F2* (1), *DPYD* (2), *G6PD* (2), *IFNL3* (1), *SLCO1B1* (2), *TPMT* (2), and *VKORC1* (1). Detailed genotyping methods are provided in the **Supplemental Methods**, and a complete list of tested variants is shown in **Table S1**.

### Medication Inclusion into Precision Medicine Analyses

The PGx analysis included 46 medications with published guidelines as of 09/25/20 by CPIC (full list available in **Table S2** and online).^5^ Drugs were considered for inclusion in the DDI analysis if they were listed as substrates, inhibitors, or inducers of CYP2B6, CYP2C19, CYP2C8, CYP2C9, CYP2D6, or CYP3A within 1) the “Clinical substrates,” “Clinical inhibitors,” or “Clinical inducers” tables of the current version (as of 09/25/20) of the U.S. Food and Drug Administration’s “Drug Development and Drug Interactions: Table of Substrates, Inhibitors and Inducers”^25^ or 2) the Indiana University School of Medicine’s Drug Interactions Flockhart Table™.^26^ Medications contained in these resources were reviewed for inclusion into DDI analyses based on the expertise of the study team. The final list of included substrates, inhibitors, and inducers are displayed in **Table S2**. Medications included in the DDI analysis between tyrosine kinase inhibitors (TKIs) and acid reducers are also listed in **Table S2**. Within our analyses, acid reducers included antacids, histamine-2 receptor antagonists (H2RAs), proton pump inhibitors (PPIs), and sucralfate.

### PGx Analyses

Within our analyses, PGx recommendations for drug-gene pairs were classified by genotype-predicted phenotype (e.g., metabolizer status) based on annotations from the Pharmacogenomics Knowledge Base (PharmGKB). Phenotypes were considered actionable if CPIC guidelines recommended a clinical action to manage the drug-gene interaction (see **File S1** for actionability determinations). Specific clinical actions included adjustment of initial or maintenance dosing, selection of alternative therapy, or performing additional tests to determine enzyme activity.

Using these determinations, we considered genotype-predicted phenotypes as actionable within our phenotype distribution (**Table 2**) if CPIC guidelines for one or more drug-gene pairs recommended clinical action based on the specified phenotype. For our analyses characterizing the prevalence of actionable PGx opportunities, we only included instances where a medication was prescribed to a subject with a CPIC-defined actionable genotype-predicted phenotype for that same medication (e.g., prescription of clopidogrel in a CYP2C19 poor metabolizer).

### DDI Analyses

DDIs involving CYP enzymes were defined as concomitant prescription of an inhibitor or inducer with a sensitive substrate of the same drug-metabolizing enzyme. To account for temporal delays in CYP induction and de-induction following the onset and offset of CYP inducers, the window for DDIs with co-administered CYP substrates was defined as starting 7 days after initiation of inducer therapy and lasting 7 days after termination of inducer therapy. DDIs involving CYP enzymes were analyzed for CYP2B6, CYP2C19, CYP2C8, CYP2C9, CYP2D6, and CYP3A for each patient from their date of first cancer diagnosis until the last date of data collection (04/20/20). DDIs were also assessed for concomitant prescription of drug-drug pairs that included TKIs and medications known to reduce gastrointestinal acidity.

Extracted medication data contained the date, time, and location (i.e., whether administered in a medical setting, including outpatient clinics, or whether dispensed from a pharmacy) for each prescription. The days supply for each prescription was conservatively estimated using the following assumptions. For prescriptions administered in a medical setting, the days supply was assumed to be one. For prescriptions dispensed from a pharmacy, the days supply was assumed based on the shortest days supply for indications for which the drug is typically prescribed (see **Table S3** for a complete list of assumed durations for all prescriptions dispensed from a pharmacy). An exception to this method was made for prescriptions dispensed from a pharmacy that were 1) dispensed for at least three consecutive regular intervals (e.g., every 30 days, every 90 days) and 2) written for medications that are commonly used as maintenance therapy for chronic medical conditions (e.g., antihypertensives). For these prescriptions, the patient was assumed to be taking the medication for the entire interval between consecutive prescriptions.

Instances of autoinhibition and autoinduction (i.e., a medication altering its own metabolism upon chronic administration) were not considered as DDIs in our analyses. In addition, DDIs involving common chemotherapy regimens (e.g., prednisone and docetaxel) were not included in our analyses. Instances of co-administration of multiple proton pump inhibitors were also not considered as DDIs.

We defined “serious DDIs” as DDI pairs with sensitive substrate drugs that have one or more of the following: 1) a narrow therapeutic index, 2) indications as cancer treatments, or 3) an association with significant adverse drug reactions (see bolded drugs in **Table S2**).

### Composite Precision Medicine Analyses

The prevalence of CYP inhibitor-mediated phenoconversion was assessed for CYP2B6, CYP2C19, CYP2C9, CYP2D6, and CYP3A4. Within our analyses, we coded subjects as positive for CYP inhibitor-mediated phenoconversion if they were (1) genotype-predicted ultrarapid, rapid, normal, or intermediate metabolizers and (2) prescribed a relevant strong CYP inhibitor at any time after first cancer diagnosis.

### Statistical Analyses

Data for the PGx and DDI analyses were analyzed using descriptive statistics (counts and percentages) using JMP Pro v.15.0.0.

## RESULTS

### Subject Demographic, Clinical, and Medication Data

Demographic and clinical characteristics of the 481 study subjects with advanced cancer included are shown in **Table 1**. Our cohort was a median of 57 ± 16.6 (median ± interquartile range [IQR]) years old, and most subjects were white (87.9%) and female (53.2%). The most common types of cancer at first diagnosis included breast (12.7%), pancreatic (10.8%), and colorectal (9.6%). The median duration of follow-up, defined as the time between the date of first cancer diagnosis and the date of last prescription, was 2.9 ± 4.9 (median ± IQR) years.

**Table 1.** Demographic and clinical characteristics of study cohort with advanced cancer.

| Variable | Value in Full Cohort (n=481) |
| --- | --- |
| Age in years at first cancer diagnosis (median [IQR]) | 57.4 (16.6) |
| Sex_(Count [Percent]) |  |
| Female | 256 (53.2%) |
| Male | 225 (46.8%) |
| Race (Count [Percent]) |  |
| White | 423 (87.9%) |
| Black | 38 (7.9%) |
| Asian | 8 (1.7%) |
| Other* | 3 (0.4%) |
| Unknown | 9 (1.9%) |
| Cancer type at first diagnosis (Count [Percent]) |  |
| Breast | 61 (12.7%) |
| Pancreatic | 52 (10.8%) |
| Colorectal | 46 (9.6%) |
| Prostate | 40 (8.3%) |
| Soft-tissue sarcoma | 36 (7.5%) |
| Ovarian | 26 (5.4%) |
| Non-small cell lung | 23 (4.8%) |
| Renal | 18 (3.7%) |
| Thymic | 13 (2.7%) |
| Cholangiocarcinoma | 12 (2.5%) |
| Head and neck | 11 (2.3%) |
| Bladder | 10 (2.1%) |
| Unknown primary | 13 (2.7%) |
| Duration of follow-up in years<sup>+</sup> (median [IQR]) | 2.9 (4.9) |

\*One individual who reported a race of “other” reported Hispanic ethnicity.
<sup>+</sup>Defined as the time elapsed between the date of first cancer diagnosis and date of most recent prescription

Extracted medication data contained ≥1 prescription for 469 out of 481 (97.5%) subjects. Filtering to include only prescriptions since each subject’s respective date of first cancer diagnosis yielded 158,188 unique prescriptions that were assessed within our precision medicine analyses (schematic of filtering results shown in **Figure S1**). Since first cancer diagnosis, our cohort had 1) a total of 7,074 unique prescriptions for medications contained within a CPIC guideline (herein called “PGx medications”) and 2) a total of 22,642 unique prescriptions for medications that were defined as inducers, inhibitors, or sensitive substrates of CYP2B6, CYP2C19, CYP2C8, CYP2C9, CYP2D6, and/or CYP3A, acid reducers, or TKIs (herein called “DDI medications”).

### PGx Analyses

The distribution of genotype-predicted phenotypes within our cohort for all pharmacogenes is displayed in **Table 2**. When defining actionable phenotypes as those with clinically actionable recommendations within CPIC guidelines for at least one medication, the rates of actionable phenotypes were highest for CYP2C19 (59.5%) and VKORC1 (52.4%) and lowest for TPMT (7.3%), G6PD (1.5%), and DPYD (1.0%).

**Table 2.** Distribution of genotype-predicted phenotypes within study cohort for major pharmacogenes.

| **Gene** | **Ultrarapid Metabolizer** | **Normal Metabolizer** | **Intermediate Metabolizer** | **Poor Metabolizer** | **Indeterminate** | **Actionable** |
| --- | --- | --- | --- | --- | --- | --- |
| *CYP2B6* |  | 248 (51.6%) | 199 (41.4%) | 34 (7.1%) |  | **233 (48.4%)** |
| *CYP2C19* | 153 (31.8%)* | 189 (39.3%) | 124 (25.8%) | 9 (1.9%) | 6 (1.2%) | **286 (59.5%)** |
| *CYP2C9* |  | 321 (66.7%) | 146 (30.4%) | 14 (2.9%) |  | **160 (33.3%)** |
| *CYP2D6* | 9 (1.9%) | 254 (52.8%) | 182 (37.8%) | 24 (5.0%) | 12 (2.5%) | **215 (44.7%)** |
| *CYP3A4* |  | 437 (90.9%) | 41 (8.5%) | 2 (0.4%) | 1 (0.2%) | **43 (8.9%)<sup>±</sup>** |
| *CYP3A5* |  | 17 (3.5%) | 71 (14.8%) | 391 (81.3%) | 2 (0.4%) | **88 (18.3%)** |
| *CYP4F2* |  | 265 (55.1%) | 175 (36.4%) | 41 (8.5%) |  | **216 (44.9%)** |
| *DPYD* |  | 476 (99.0%) | 5 (1.0%) | 0 (0%) |  | **5 (1.0%)** |
| *G6PD*<sup>+</sup> |  | 474 (98.5%) | 4 (0.8%) | 3 (0.6%) |  | **7 (1.5%)** |
| *IFNL3 (IL28B)*<sup>+</sup> |  | 204 (42.4%) | 220 (45.7%) | 57 (11.9%) |  | **0 (0%)** |
| *SLCO1B1*<sup>+</sup> |  | 333 (69.2%) | 107 (22.2%) | 11 (2.3%) | 30 (6.2%) | **118 (24.5%)** |
| *TPMT* |  | 440 (91.5%) | 35 (7.3%) | 0 (0%) | 6 (1.2%) | **35 (7.3%)** |
| *VKORC1*<sup>+</sup> |  | 229 (47.6%) | 199 (41.4%) | 53 (11.0%) |  | **252 (52.4%)** |

\*For CYP2C19, count in ultrarapid metabolizer column includes counts of both ultrarapid metabolizers (n=20) and rapid metabolizers (n=133).
<sup>+</sup>For designated genes, “normal metabolizer,” “intermediate metabolizer,” and “poor metabolizer” designations refer to subjects who are non-carriers, heterozygous, and homozygous for CPIC-defined actionable variants, respectively.
<sup>±</sup>While CPIC does not make *CYP3A4* genetic-guided recommendations for any drugs, we classify subjects with one or two copies of the *CYP3A4*\*22 loss-of-function allele as intermediate and poor metabolizers, respectively, and consider these phenotypes to be actionable since they are used at our institution to guide tacrolimus dosing in CYP3A5 non-expressers.

Of 469 analyzed subjects, 282 (60.1%) were prescribed at least one PGx medication. These included a total of 1,045 unique PGx medications (i.e., prescription of a unique PGx medication for a unique subject), with an average of 2.2 ± 2.4 (mean ± standard deviation) PGx medications/subject and a maximum of 12 PGx medications in one subject. When considering both prescribed medications and genotype-predicted phenotypes, we identified a total of 81 unique opportunities for “actionable PGx,” defined as an instance where a PGx medication was prescribed to a subject with an actionable phenotype based on CPIC recommendations. Instances of actionable PGx occurred for 67 subjects (14.3%), with 56 subjects having instances of actionable PGx involving 1 medication, 8 subjects having actionable PGx involving 2 medications, and 3 subjects having actionable PGx involving 3 medications.

The prevalence of instances of actionable PGx, when stratified by the drug-gene pairs involved, are shown in **Table 3**. For PGx medications prescribed in at least five subjects, the rates of actionable PGx were highest for warfarin (87.5%), amitriptyline (58.3%), and clopidogrel (42.9%). Conversely, capecitabine, 5-fluorouracil, sertraline, and celecoxib had no instances for actionable PGx. For warfarin, subjects had actionable PGx recommendations based on *CYP2C9, CYP4F2*, and *VKORC1* genotype-based phenotypes in 20.8%, 58.3%, and 50.0% of cases, respectively. For amitriptyline, subjects had actionable PGx recommendations based on *CYP2C19* and *CYP2D6* genotype-based phenotypes in 16.7% and 50.0% of cases, respectively.

**Table 3.** Prevalence of PGx medications prescribed in subjects with clinically actionable genotype-predicted phenotypes based on CPIC recommendations.

| **Drug-Gene Pair** | **# Prescribed Drug** | **% with Actionable PGx** |
| --- | --- | --- |
| Ondansetron-*CYP2D6* | 256 | 0.8% |
| Pantoprazole-*CYP2C19* | 171 | 4.7% |
| Omeprazole-*CYP2C19* | 99 | 2.0% |
| Ibuprofen-*CYP2C9* | 93 | 5.4% |
| Tramadol-*CYP2D6* | 81 | 6.2% |
| Capecitabine-*DPYD* | 62 | 0% |
| 5-Fluorouracil-*DPYD* | 40 | 0% |
| Sertraline-*CYP2C19* | 28 | 0% |
| Escitalopram-*CYP2C19* | 25 | 12.0% |
| Lansoprazole-*CYP2C19* | 24 | 4.2% |
| Warfarin-*CYP2C9/CYP4F2/VKORC1* | 24 | 87.5% |
| Citalopram-*CYP2C19* | 21 | 23.8% |
| Simvastatin-*SLCO1B1* | 21 | 23.8% |
| Meloxicam-*CYP2C9* | 16 | 12.5% |
| Celecoxib-*CYP2C9* | 15 | 0% |
| Amitriptyline-*CYP2C19/CYP2D6* | 12 | 58.3% |
| Nortriptyline-*CYP2D6* | 9 | 11.1% |
| Dexlansoprazole-*CYP2C19* | 8 | 12.5% |
| Clopidogrel-*CYP2C19* | 7 | 42.9% |
| Codeine-*CYP2D6* | 7 | 14.3% |
| Paroxetine-*CYP2D6* | 6 | 16.7% |
| Tamoxifen-*CYP2D6* | 6 | 33.3% |
| Doxepin-*CYP2C19/CYP2D6* | 3 | 100% |
| Voriconazole-*CYP2C19* | 3 | 66.7% |
| Rasburicase-*G6PD* | 2 | 0% |
| Atomoxetine-*CYP2D6* | 1 | 0% |
| Azathioprine-*TPMT* | 1 | 0% |
| Imipramine-*CYP2C19/CYP2D6* | 1 | 100% |
| Phenytoin-*CYP2C9* | 1 | 0% |
| Ribavirin-*IFNL3* | 1 | 0% |
| Tacrolimus-*CYP3A5* | 1 | 0% |
| **Total** | **1045** |  |

The “number prescribed drug” indicates the number of subjects within our cohort that were prescribed the corresponding drug. The “percent with actionable PGx,” which was calculated at the subject-level, indicates the percent of subjects prescribed the corresponding drug that had genotypes for which current CPIC guidelines recommend actionable clinical management strategies.

### DDI Analyses

Of 469 analyzed subjects, the prevalence of ≥1 prescription for an inducer, inhibitor, or substrate of any CYP enzyme was 49.0%, 58.0%, and 64.0%, respectively. **Figure S2** displays the prevalence of subjects with prescriptions for inducers, inhibitors, and substrates across the six enzyme systems that were assessed. Prescriptions for CYP inducers were most common for CYP2C19, CYP2C9, and CYP3A, occurring in 49.0% of subjects. Prescriptions for inhibitors were most common for CYP2D6 (occurring in 53.3% of subjects), CYP2C9 (35.0%), CYP3A (33.9%), and CYP2C19 (31.8%), while prescriptions for sensitive substrates were most common for CYP3A, CYP2D6, and CYP2C19 (prescribed in 60.3%, 59.9%, and 48.2% of subjects, respectively).

When assessing concomitant prescription of both a relevant perpetrator (inducer or inhibitor) and victim (sensitive substrate) drug, 236 subjects (50.3%) had a DDI affecting at least one CYP enzyme system. Given the frequent use of corticosteroids to treat and manage treatment-related complications for many types of cancer,^27^ we also performed DDI analyses excluding corticosteroids, which are potent inducers of CYP2C19, CYP2C9, and CYP3A; 225 subjects (48.0%) had a DDI affecting at least one major CYP enzyme when excluding corticosteroids. As shown in **Table 4**, the prevalence of DDIs in our cohort was highest for CYP2D6 (affecting 45.2% of subjects; average of 1.5 DDIs/subject), followed by CYP3A (29.9%; 0.8 DDIs/subject), CYP2C19 (23.9%; 0.5 DDIs/subject), CYP2C9 (11.7%; 0.2 DDIs/subject), CYP2B6 (0.2%), and CYP2C8 (0%). When excluding corticosteroids, the prevalence of DDIs for CYP2C19, CYP2C9, and CYP3A was reduced to 10.2%, 7.0%, and 20.3%, respectively (**Table 4**). The most common drug-drug pairs contained within observed DDIs, stratified by enzyme, are shown in **Table S4**. The subject-level prevalence for serious DDIs, which were classified by the substrates involved, was 34.8% for any CYP enzyme when including corticosteroids and 29.4% when excluding corticosteroids (**Table 4**). Serious DDIs were most common for CYP3A, occurring in 24.9% of subjects and including sensitive substrates like fentanyl, midazolam, and tramadol. In contrast, serious DDIs were less common for CYP2C19 (11.7% of subjects; sensitive substrates included escitalopram, sertraline, and citalopram), CYP2C9 (4.7% of subject; substrates included warfarin, dronabinol, and phenytoin), and CYP2D6 (16.8% of subject; substrates included tramadol, sertraline, and mirtazapine). When adjusting the prevalence of CYP enzyme-mediated DDIs based on subject genotype (i.e., excluding DDIs involving inducer or inhibitor drugs in subjects who are genotype-predicted poor metabolizers), the subject-level prevalence is as follows: CYP2B6: 0.2%; CYP2C19: 23.9%; CYP2C8: 0%; CYP2C9: 11.7%; CYP2D6: 44.1%; and CYP3A: 29.6% (adjusted based on *CYP3A4* genotype).

**Table 4.** Number and prevalence of unique DDIs (i.e., unique co-prescription of a relevant drug-drug pair in a unique subject) by enzyme involved in n=469 subjects prescribed ≥ 1 medication, including (left) and excluding (right) DDIs involving corticosteroids.

| Enzyme | DDIs Including Corticosteroids |  |  |  | DDIs Excluding Corticosteroids |  |  |  |
| --- | --- | --- | --- | --- | --- | --- | --- | --- |
|  | Total DDIs | DDIs/Subject (Mean) | DDI Prevalence (%) | Serious DDI Prevalence (%) | Total DDIs | DDIs/Subject (Mean) | DDI Prevalence (%) | Serious DDI Prevalence (%) |
| CYP2B6 | 1 | 0.00 | 0.2% | 0.2% | 1 | 0.00 | 0.2% | 0.2% |
| CYP2C19 | 237 | 0.51 | 23.9% | 11.7% | 89 | 0.19 | 10.2% | 5.8% |
| CYP2C8 | 0 | 0 | 0% | 0% | 0 | 0 | 0% | 0% |
| CYP2C9 | 76 | 0.16 | 11.7% | 4.7% | 39 | 0.08 | 7.0% | 2.3% |
| CYP2D6 | 695 | 1.48 | 45.2% | 16.8% | 695 | 1.48 | 45.2% | 16.8% |
| CYP3A | 392 | 0.84 | 29.9% | 24.9% | 217 | 0.46 | 20.3% | 18.6% |
| **Any DDI** | **1401** | **2.99** | **50.3%** | **34.8%** | **1041** | **2.22** | **48.0%** | **29.4%** |

**Note:** All DDI prevalence calculations are at the subject level.

TKIs have emerged as first-line treatment options for a variety of cancers. However, multiple investigations have described the potential for significant DDIs involving orally-administered TKIs and acid reducing agents, including antacids, H2RAs, and PPIs, that reduce TKI bioavailability and impact treatment outcomes.^28-31^ Accordingly, we characterized the prevalence of DDIs involving TKIs and acid reducers in our study population. Within our cohort, 68 subjects (14.5%) were prescribed at least one TKI, with pazopanib (prescribed in 17 subjects), sunitinib (10), and crizotinib (9) being the most commonly prescribed. Of the 68 subjects prescribed a TKI, 33 (48.5%) had a concomitant prescription of at least one acid reducer. Within our population, the most common acid reducer classes involved in DDIs were PPIs (perpetrator drug in 34 DDIs), followed by H2RAs (10) and antacids (6).

### Composite Precision Medicine Analyses

To assess the prevalence of composite opportunities for precision medicine interventions, we aggregated findings from our actionable PGx, serious CYP-mediated DDI, and acid reducer-TKI DDI analyses at the subject level. As shown in **Figure 1**, 186 subjects (39.7%) had at least one opportunity for a precision medicine intervention. 68 subjects (14.5%) had opportunities for more than one type of precision medicine intervention, with 9 of these subjects (1.9%) having opportunities for PGx and management of both CYP-mediated and acid reducer-TKI DDIs.

**Figure 1.**
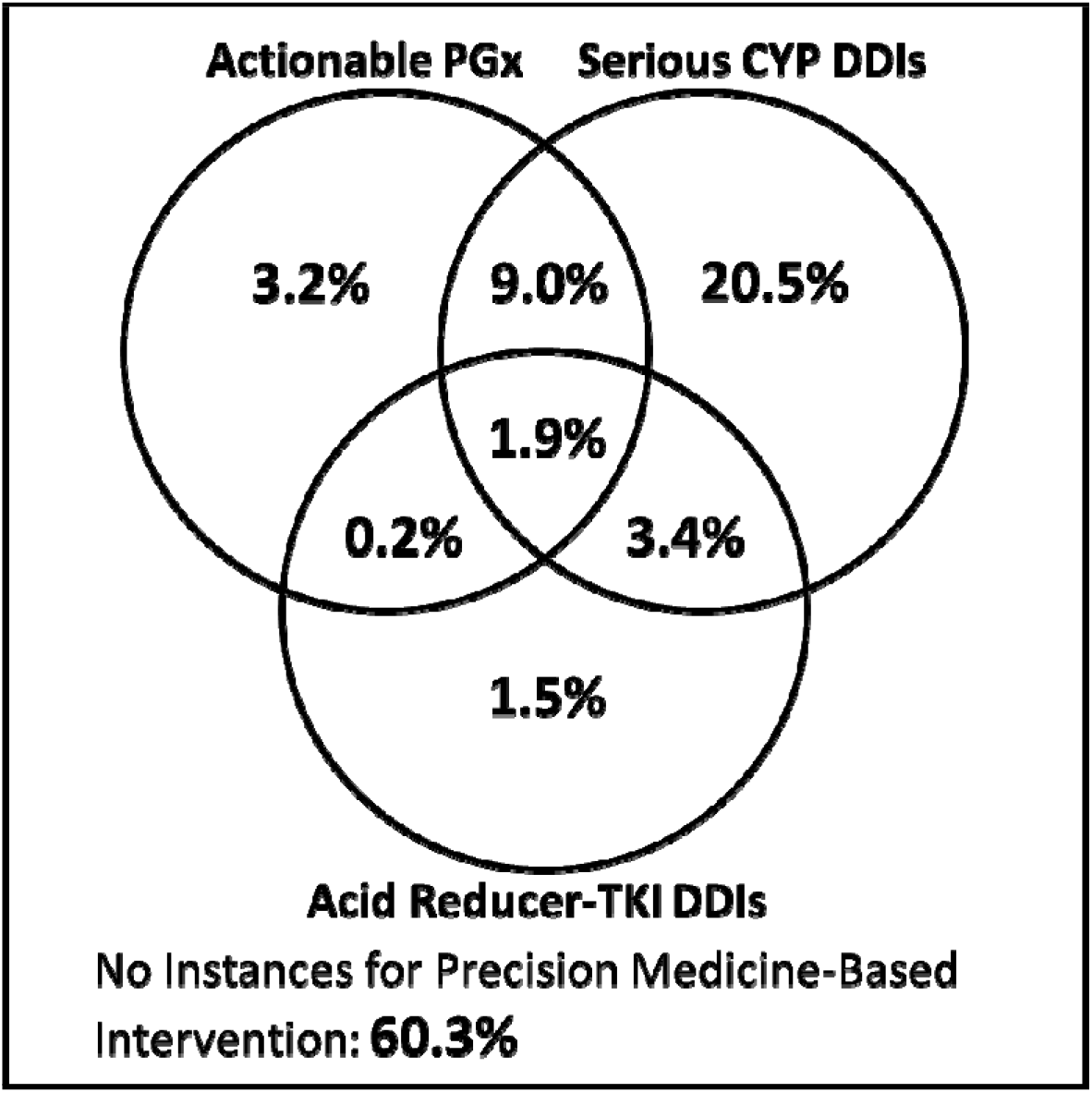
Subject-level prevalence for composite precision medicine opportunities, including actionable PGx, management of serious CYP-mediated DDIs, and management of DDIs including acid reducers and TKIs.

Finally, we assessed the prevalence of CYP inhibitor-mediated phenoconversion, the process by which co-administration of a strong inhibitor functionally converts those with any genotype to a poor metabolizer phenotype, for CYP2B6, CYP2C19, CYP2C9, CYP2D6, and CYP3A4. As shown in **Figure 2**, CYP inhibitor-mediated phenoconversion enhanced the number of subjects with actionable phenotypes for CYP2C19, CYP2C9, and CYP2D6, and CYP3A4, increasing the prevalence from 59.5% to 72.8%, 33.3% to 55.9%, 44.7% to 76.3%, and 8.9% to 38.9%, respectively. In contrast, CYP inhibitor-mediated phenoconversion only slightly changed the number of actionable phenotypes for CYP2B6 (prevalence increased from 48.4% to 49.1%) due to the low prevalence of prescription of CYP2B6 inhibitors within our cohort. When considering all five investigated CYPs together, nearly every subject in our cohort (98.3%) had an actionable phenotype (either genotype-predicted or from CYP inhibitor-mediated phenoconversion) for at least one CYP since their date of first cancer diagnosis. Also, 47 subjects (9.8%) had genotype-predicted or phenoconverted actionable phenotypes for all five CYP enzymes.

**Figure 2.**
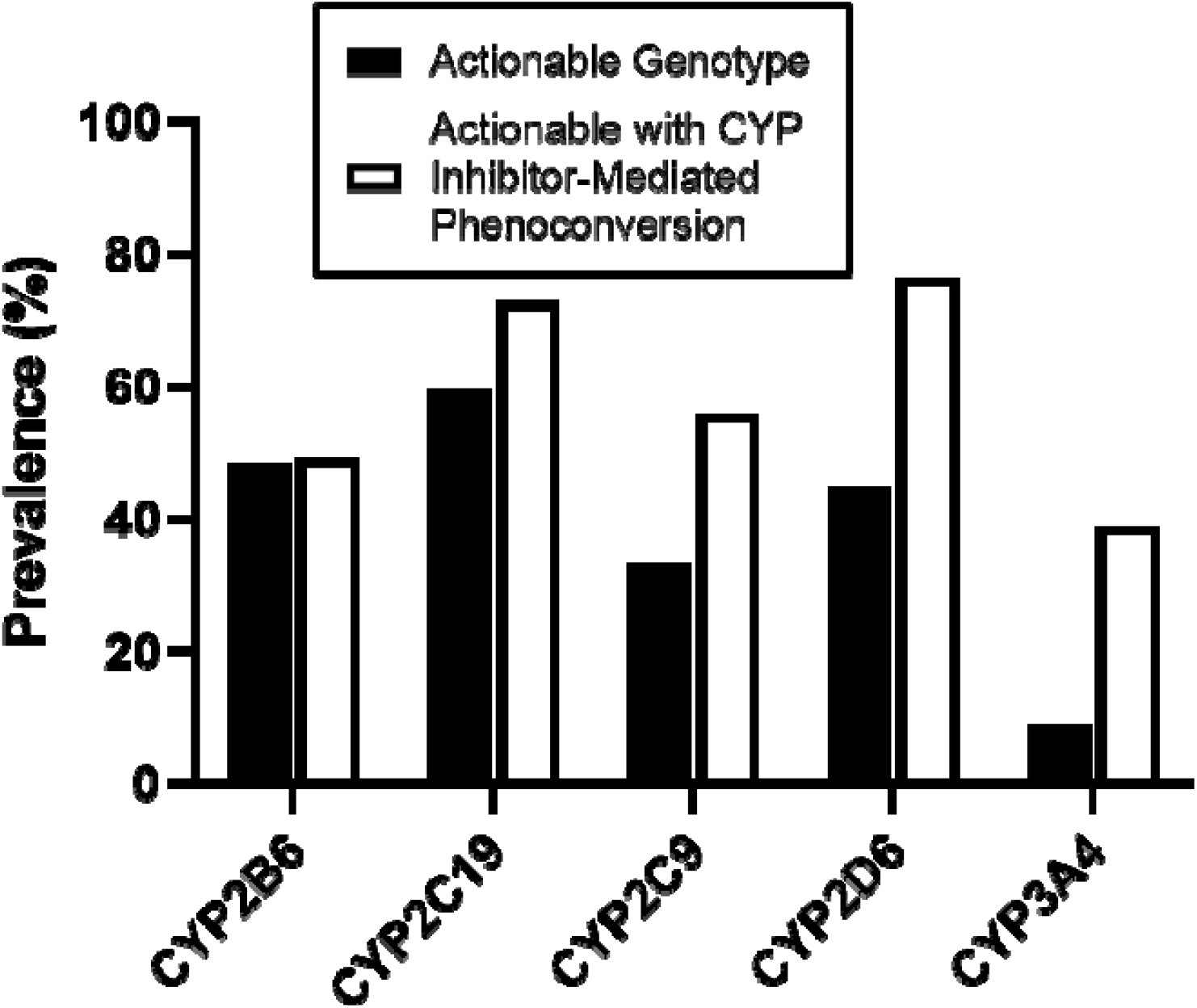
Subject-level prevalence of clinically actionable phenotypes for major CYP enzymes based on genotype and due to CYP inhibitor-mediated phenoconversion.

## DISCUSSION

In this investigation, we provide quantitative evidence to support the immense clinical opportunities for precision medicine approaches, including germline PGx and management of DDIs, in a cohort of patients with advanced cancer. Our findings indicate that ∼14% of subjects had opportunities for actionable PGx (i.e., prescription of a PGx medication to a subject with a CPIC guideline-defined actionable genotype) and that ∼35% and ∼7% of subjects had a serious DDI involving major CYP enzymes and acid reducers co-prescribed with TKIs, respectively. When incorporating both PGx and DDIs, we found that ∼40% of subjects had at least one opportunity for a precision medicine-based intervention and nearly all subjects (∼98%) had an actionable phenotype (genetically-predicted or drug-induced) for ≥1 CYP enzyme. Based on our findings, implementation of precision medicine approaches at first cancer diagnosis is likely to provide clinical benefit to a significant proportion of patients. Although a limited number of other studies have addressed similar topics, our investigation has significant methodological advantages, including 1) a larger cohort (n=481), 2) a broader PGx analysis consisting of 13 CPIC-actionable pharmacogenes, and 3) utilization of a statewide data repository to enable more comprehensive collection of medication data.

Previous investigations have demonstrated the potential clinical impact of PGx approaches in patients with advanced cancer. Nichols, et al. catalogued medications in a cohort of 193 patients with advanced cancer, demonstrating that 65% of patients were taking at least one PGx medication (i.e., those with a CPIC guideline).^32^ Using population estimates of allele frequencies, the authors predicted that 7.1% of patients in their cohort could benefit from at least one PGx intervention involving medications associated with nine major pharmacogenes: *CYP2C19, CYP2C9, CYP2D6, CYP3A5, CYP4F2, DPYD, HLA-B, SLCO1B1, VKORC1*. Similarly, Hertz, et al. found that 2.6% of 115 adult and pediatric patients with cancer could have benefitted from a PGx intervention involving substrates of their analyzed drug-metabolizing enzymes, which included CYP2C19, DPYD, and TPMT.^33^ An investigation by Kasi, et al. also predicted abundant opportunities for PGx interventions within their cohort of 155 patients with advanced cancer based on patient genotypes for major CYP450 enzymes, though they did not specifically collect and analyze medication data.^34^ Many of our findings are similar to those reported in past investigations. For instance, our findings related to the prevalence of prescription of PGx medications are remarkably similar to results from Nichols, et al. when considering both prescription of any PGx medication (∼60% in our analysis vs. 65% in their study) and prescription of specific PGx drugs such as ondansetron, capecitabine, and simvastatin.^32^ Our findings related to the distribution of actionable phenotypes are also consistent with those from past investigations^32,34^ as well as those predicted from large analyses of population allele frequencies.^35^ In contrast, our finding for the prevalence of subjects with potential PGx interventions (14.3%) is higher than those reported by Nichols, et al. (7.1%) or Hertz, et al. (2.6%).^32,33^ These differences are likely attributable to the facts that we (1) investigated the potential for PGx interventions across a wider array of pharmacogenes and (2) that we utilized a statewide repository with prescription data from 38 health systems to enhance the richness of our collected medication data.

Multiple investigations have also characterized the clinical potential of DDI management strategies in adult patients with advanced cancer. A 2009 review by Riechelmann, et al. summarized the prevalence of potential DDIs from six studies, finding rates between 27% and 72%.^36^ The high variability they observed among studies is likely attributable to differences in employed methodologies (e.g., utilizing patient-verified medication lists versus all drugs listed in EHRs, focusing on all potential drug interactions versus only those involving cancer medications). More recently, investigations within the U.S. and abroad have characterized the prevalence of potential DDIs in cancer patients, finding rates of 40-78%.^37-41^ Again, it appears that observed differences in potential DDI prevalence are due to methodological differences among the studies. For instance, we found that studies that included DDIs based on both pharmacokinetic (i.e., concomitant administration of an inhibitor or inducer of a drug-metabolizing enzyme along with a sensitive substrate of that enzyme) and pharmacodynamic (i.e., concomitant administration of two or more drugs with the same adverse event profile) mechanisms had higher rates of potential DDIs.^38,39^ Similarly, studies that utilized medication lists taken from the EHR (rather than those verified by patients during medication reconciliation) had higher potential DDI prevalence.^38,39^ The overall DDI prevalence of ∼52% in our study falls in the middle of those reported by past investigations. In terms of methodology, extracting medication data from the EHR likely resulted in a higher DDI prevalence in our study relative to those that used patient-verified medications. We attempted to control for this by utilizing prescription dates to only identify potential DDIs when there was temporal overlap in the prescription (and presumed coadministration) of perpetrator and victim drugs for the same CYP enzyme. Relative to other studies, our DDI prevalence was likely more conservative based on other elements in our methodology, including (1) that we excluded DDIs with pharmacodynamic mechanisms and (2) that we excluded DDIs involving drugs commonly co-administered as cancer treatment regimens (e.g., corticosteroids co-administered with docetaxel or vincristine). Our rationale for excluding these DDIs was that, in the case of pharmacodynamic DDIs, co-administration of drugs with similar adverse event profiles is often clinically indicated (e.g., dual antiplatelet therapy) and, in the case of DDIs within established cancer regimens, treating clinicians are familiar with these DDIs and have likely already determined a favorable risk-benefit ratio for the patient before prescribing. Therefore, based on these methodological elements, we believe our findings represent a conservative estimate of the prevalence of potential DDIs in advanced cancer patients involving major CYP enzymes. Additionally, our study expands on past investigations assessing potential DDI prevalence in a few significant ways. First, our results stratified DDI prevalence by the CYP enzymes involved, which could aid clinicians in selecting drugs with metabolic pathways less likely to be associated with DDIs. Next, we specifically investigated the prevalence of DDIs for acid reducing agents and TKIs, which has emerged as an important consideration in cancer precision medicine.^42^ Finally, we performed both a composite subject-level analysis and CYP inhibitor-mediated phenoconversion analysis to elucidate the net prevalence of precision medicine opportunities in our study cohort that incorporate both PGx and DDI management approaches.

Our findings are impactful since they demonstrate the abundant clinical opportunities for precision medicine approaches to optimize medication therapy in patients with advanced cancer. Specifically, we found that ∼60% of subjects in our cohort were prescribed at least one PGx medication and that approximately 1 in 7 subjects had an opportunity for actionable PGx since their date of first cancer diagnosis. These findings directly support the clinical utility of PGx approaches in patients with cancer, including the suggestion of preemptive genotyping at first cancer diagnosis.^22^ Advances in technology have also improved the feasibility of PGx approaches by reducing the costs associated with obtaining genetic information and enabling repurposing of genetic information obtained from molecular tumor boards.^24^ In addition, economic analyses have demonstrated cost savings due to toxicity sparing for both *DPYD* and *TPMT* testing.^43,44^ As a result, there is clinical momentum for standardized testing of PGx markers associated with fluoropyrimidine and thiopurine chemotherapies.^45,46^ Our findings also corroborate those from other studies^32,34^ in identifying significant opportunities for PGx to optimize supportive care therapies in patients with cancer, including SSRIs, TCAs, opioids, and commonly used antiemetics (e.g., ondansetron), based on CPIC guidelines.^19-21,47^

Related to the clinical opportunities for DDI management strategies, we found that slightly over half of our study subjects had a DDI affecting at least one major CYP enzyme since first cancer diagnosis. This finding is important given that DDIs have been associated with poor clinical outcomes and increased adverse drug events in cancer patients. For instance, CYP-mediated DDIs have been shown to increase the rates of adverse events attributable to both cancer therapies (e.g., increased paclitaxel-induced peripheral neuropathy during co-treatment with clopidogrel)^48^ and concomitant medications (e.g., increased warfarin-induced bleeding during co-treatment with capecitabine)^49^ in patients with cancer. Additionally, several studies have investigated the potential for DDIs between acid reducing agents and TKIs, demonstrating reduced progression-free and overall survival during concomitant therapy attributed to reduced TKI systemic absorption.^28-30^ Our findings support the potential for clinically significant DDIs between acid reducers and TKIs since we observed that these DDIs occurred in nearly half of subjects that were prescribed a TKI. However, it is possible that the providers told the patient to discontinue the acid reducers while taking the TKI’s. Nonetheless, our findings support the clinical potential of DDI management strategies, which have been shown to improve outcomes in other populations,^9^ in patients with advanced cancer. Finally, our work serves as one of the first investigations to assess the prevalence of potential drug-drug-gene interactions (DDGIs) (i.e., CYP inhibitor-mediated phenoconversion) within a clinical cohort. While the strategies to manage DDGIs borrow from both PGx and DDI management approaches, consideration of DDGIs may provide critical information that modifies the risk of adverse drug events predicted from consideration of either approach in isolation.^50^ As demonstrated by our composite study findings that ∼40% of subjects had at least one opportunity for precision medicine intervention and ∼98% of subjects had an actionable phenotype for ≥1 CYP enzymes, PGx information and concomitant drug lists should be used in tandem to most accurately inform approaches to optimize medication therapy. Given the complexities of DDGIs, including concepts like phenoconversion and interplay of multiple drug biotransformation pathways, expert guidance that includes perspectives from both clinical pharmacologists and clinicians is needed to inform actionable clinical management strategies.

We acknowledge several limitations of our study. First, our extracted medication data did not include a way to conclusively ascertain days supply in order to assess temporal overlap between perpetrator and victim drugs within our DDI analyses. To compensate for this limitation, we used conservative methods in our DDI analyses to estimate days supply for each prescription, as described in the methods. While this limitation may have influenced our findings related to the prevalence of DDIs, it did not impact results from our PGx analyses. Our extracted medication data also did not consistently contain information about the medication dose. As a result, our analysis may have overestimated the prevalence of instances for actionable PGx with amitriptyline since current CPIC guidelines do not recommend clinical action at daily doses under 50 mg.^19^ Also, our panel-based genotyping method only tested for relatively common functional variants in the assessed genes within our primary ethnic and racial populations, potentially excluding rare functional variants that alter drug response. While we do not expect that this approach significantly impacted our findings, it is important to note that utilizing panel-based genotyping (as opposed to a more complete approach like whole genome sequencing) may have caused us to underestimate the actual clinical opportunities for actionable PGx in our cohort. Additionally, advances in knowledge since study initiation limited our ability to assess variants with newly established relevance to pharmacotherapy (e.g., HapB3 in *DPYD*).Our genotyping panel also did not assess every pharmacogene included within a CPIC guideline. However, the pharmacogenes covered in our panel serve as the genetic basis for over 80% of the PGx recommendations contained within current CPIC guidelines.^5^

In conclusion, our work provides quantitative evidence of the vast clinical opportunities for precision medicine approaches in patients with advanced cancer, demonstrating the clinical utility of both germline PGx and DDI management strategies. Given their established clinical benefits and the abundant opportunities for their use demonstrated by our results, precision medicine approaches are likely to improve medication outcomes in cancer patients and may provide clinical benefit if incorporated into the workflow of molecular tumor boards. In order to facilitate widespread adoption of precision medicine approaches in this high-value patient population, future research is needed to (1) prospectively demonstrate the clinical benefit of precision medicine approaches on patient outcomes and to (2) identify effective strategies for clinical implementation of precision medicine approaches.

## Data Availability

Data are available at the request of the corresponding author.

## ACKNOWLEDGEMENTS

None

## REFERENCES

1. Claassens DMF, Vos GJA, Bergmeijer TO, et al: A Genotype-Guided Strategy for Oral P2Y(12) Inhibitors in Primary PCI. N Engl J Med 381:1621–1631, 2019

2. Mallal S, Phillips E, Carosi G, et al: HLA-B*5701 screening for hypersensitivity to abacavir. N Engl J Med 358:568–79, 2008

3. Pirmohamed M, Burnside G, Eriksson N, et al: A randomized trial of genotype-guided dosing of warfarin. N Engl J Med 369:2294–303, 2013

4. U.S. Food and Drug Administration: Table of Pharmacogenomic Biomarkers in Drug Labeling, (ed 08/18/20), 2020

5. Clinical Pharmacogenetics Implementation Consortium: Guidelines, 2020

6. Shugg T, Pasternak AL, London B, et al: Prevalence and types of inconsistencies in clinical pharmacogenetic recommendations among major U.S. sources. NPJ Genom Med 5:48, 2020

7. Bates DW, Cullen DJ, Laird N, et al: Incidence of adverse drug events and potential adverse drug events. Implications for prevention. ADE Prevention Study Group. Jama 274:29–34, 1995

8. Wright A, Feblowitz J, Phansalkar S, et al: Preventability of adverse drug events involving multiple drugs using publicly available clinical decision support tools. Am J Health Syst Pharm 69:221–7, 2012

9. Arnold RJG, Tang J, Schrecker J, et al: Impact of Definitive Drug-Drug Interaction Testing on Medication Management and Patient Care. Drugs Real World Outcomes 5:217–224, 2018

10. U.S. Food and Drug Administration: Clinical Drug Interaction Studies -- Cytochrome P450 Enzyme-and Transporter-Mediated Drug Interactions: Guidance for Industry, (ed January 2020), 2020

11. Department of Health and Human Services Office of AIDS Research Advisory Council: Guidelines for the Use of Antiretroviral Agents in Adults and Adolescents with HIV, 2018

12. Grundy SM, Stone NJ, Bailey AL, et al: 2018 AHA/ACC/AACVPR/AAPA/ABC/ACPM/ADA/AGS/APhA/ASPC/NLA/PCNA Guideline on the Management of Blood Cholesterol: A Report of the American College of Cardiology/American Heart Association Task Force on Clinical Practice Guidelines. Circulation 139:e1082–e1143, 2019

13. LeBlanc TW, McNeil MJ, Kamal AH, et al: Polypharmacy in patients with advanced cancer and the role of medication discontinuation. Lancet Oncol 16:e333–41, 2015

14. Guthrie B, Makubate B, Hernandez-Santiago V, et al: The rising tide of polypharmacy and drug-drug interactions: population database analysis 1995-2010. BMC Med 13:74, 2015

15. Sharma M, Vadhariya A, Chikermane S, et al: Clinical Outcomes Associated with Drug-Drug Interactions of Oral Chemotherapeutic Agents: A Comprehensive Evidence-Based Literature Review. Drugs Aging 36:341–354, 2019

16. Kato S, Kim KH, Lim HJ, et al: Real-world data from a molecular tumor board demonstrates improved outcomes with a precision N-of-One strategy. Nat Commun 11:4965, 2020

17. Relling MV, Schwab M, Whirl-Carrillo M, et al: Clinical Pharmacogenetics Implementation Consortium Guideline for Thiopurine Dosing Based on TPMT and NUDT15 Genotypes: 2018 Update. Clin Pharmacol Ther 105:1095–1105, 2019

18. Amstutz U, Henricks LM, Offer SM, et al: Clinical Pharmacogenetics Implementation Consortium (CPIC) Guideline for Dihydropyrimidine Dehydrogenase Genotype and Fluoropyrimidine Dosing: 2017 Update. Clin Pharmacol Ther 103:210–216, 2018

19. Hicks JK, Sangkuhl K, Swen JJ, et al: Clinical pharmacogenetics implementation consortium guideline (CPIC) for CYP2D6 and CYP2C19 genotypes and dosing of tricyclic antidepressants: 2016 update. Clin Pharmacol Ther 102:37–44, 2017

20. Crews KR, Gaedigk A, Dunnenberger HM, et al: Clinical Pharmacogenetics Implementation Consortium guidelines for cytochrome P450 2D6 genotype and codeine therapy: 2014 update. Clin Pharmacol Ther 95:376–82, 2014

21. Hicks JK, Bishop JR, Sangkuhl K, et al: Clinical Pharmacogenetics Implementation Consortium (CPIC) Guideline for CYP2D6 and CYP2C19 Genotypes and Dosing of Selective Serotonin Reuptake Inhibitors. Clin Pharmacol Ther 98:127–34, 2015

22. Hoffman JM, Haidar CE, Wilkinson MR, et al: PG4KDS: a model for the clinical implementation of pre-emptive pharmacogenetics. Am J Med Genet C Semin Med Genet 166c:45–55, 2014

23. van der Lee M, Allard WG, Bollen S, et al: Repurposing of Diagnostic Whole Exome Sequencing Data of 1,583 Individuals for Clinical Pharmacogenetics. Clin Pharmacol Ther 107:617–627, 2020

24. Numanagić I, Malikić S, Ford M, et al: Allelic decomposition and exact genotyping of highly polymorphic and structurally variant genes. Nat Commun 9:828, 2018

25. U.S. Food and Drug Administration: Drug Development and Drug Interactions: Table of Substrates, Inhibitors and Inducers, (ed 03/10/20), 2020

26. Indiana University School of Medicine: Drug Interactions Flockhart Table, 2020

27. Lossignol D: A little help from steroids in oncology. J Transl Int Med 4:52–54, 2016

28. Chen YM, Lai CH, Chang HC, et al: Antacid Use and De Novo Brain Metastases in Patients with Epidermal Growth Factor Receptor-Mutant Non-Small Cell Lung Cancer Who Were Treated Using First-Line First-Generation Epidermal Growth Factor Receptor Tyrosine Kinase Inhibitors. PLoS One 11:e0149722, 2016

29. Chu MP, Ghosh S, Chambers CR, et al: Gastric Acid suppression is associated with decreased erlotinib efficacy in non-small-cell lung cancer. Clin Lung Cancer 16:33–9, 2015

30. Ha VH, Ngo M, Chu MP, et al: Does gastric acid suppression affect sunitinib efficacy in patients with advanced or metastatic renal cell cancer? J Oncol Pharm Pract 21:194–200, 2015

31. Mir O, Touati N, Lia M, et al: Impact of Concomitant Administration of Gastric Acid-Suppressive Agents and Pazopanib on Outcomes in Soft-Tissue Sarcoma Patients Treated within the EORTC 62043/62072 Trials. Clin Cancer Res 25:1479–1485, 2019

32. Nichols D, Arnold S, Weiss HL, et al: Pharmacogenomic potential in advanced cancer patients. Am J Health Syst Pharm 76:415–423, 2019

33. Hertz DL, Glatz A, Pasternak AL, et al: Integration of Germline Pharmacogenetics Into a Tumor Sequencing Program. JCO Precis Oncol 2, 2018

34. Kasi PM, Koep T, Schnettler E, et al: Feasibility of Integrating Panel-Based Pharmacogenomics Testing for Chemotherapy and Supportive Care in Patients With Colorectal Cancer. Technol Cancer Res Treat 18:1533033819873924, 2019

35. Zhou Y, Ingelman-Sundberg M, Lauschke VM: Worldwide Distribution of Cytochrome P450 Alleles: A Meta-analysis of Population-scale Sequencing Projects. Clin Pharmacol Ther 102:688–700, 2017

36. Riechelmann RP, Del Giglio A: Drug interactions in oncology: how common are they? Ann Oncol 20:1907–12, 2009

37. Chen L, Cheung WY: Potential drug interactions in patients with a history of cancer. Curr Oncol 21:e212–20, 2014

38. Ismail M, Khan S, Khan F, et al: Prevalence and significance of potential drug-drug interactions among cancer patients receiving chemotherapy. BMC Cancer 20:335, 2020

39. Korucu FC, Senyigit E, Köstek O, et al: A retrospective study on potential drug interactions: A single center experience. Journal of Oncological Sciences 4:80–84, 2018

40. van Leeuwen RW, Brundel DH, Neef C, et al: Prevalence of potential drug-drug interactions in cancer patients treated with oral anticancer drugs. Br J Cancer 108:1071–8, 2013

41. van Leeuwen RW, Swart EL, Boven E, et al: Potential drug interactions in cancer therapy: a prevalence study using an advanced screening method. Ann Oncol 22:2334–41, 2011

42. Yu G, Zheng QS, Wang DX, et al: Drug interactions between tyrosine-kinase inhibitors and acid suppressive agents: more than meets the eye. Lancet Oncol 15:e469–70, 2014

43. Zarca K, Durand-Zaleski I, Loriot MA, et al: Modeling the Outcome of Systematic TPMT Genotyping or Phenotyping Before Azathioprine Prescription: A Cost-Effectiveness Analysis. Mol Diagn Ther 23:429–438, 2019

44. Fragoulakis V, Roncato R, Fratte CD, et al: Estimating the Effectiveness of DPYD Genotyping in Italian Individuals Suffering from Cancer Based on the Cost of Chemotherapy-Induced Toxicity. Am J Hum Genet 104:1158–1168, 2019

45. Weitzel KW, Smith DM, Elsey AR, et al: Implementation of Standardized Clinical Processes for TPMT Testing in a Diverse Multidisciplinary Population: Challenges and Lessons Learned. Clin Transl Sci 11:175–181, 2018

46. Hertz DL, Sahai V: Including DPYD on Cancer Genetic Panels to Prevent Fatal Fluoropyrimidine Toxicity. J Natl Compr Canc Netw 18:372–374, 2020

47. Bell GC, Caudle KE, Whirl-Carrillo M, et al: Clinical Pharmacogenetics Implementation Consortium (CPIC) guideline for CYP2D6 genotype and use of ondansetron and tropisetron. Clin Pharmacol Ther 102:213–218, 2017

48. Agergaard K, Mau-Sørensen M, Stage TB, et al: Clopidogrel-Paclitaxel Drug-Drug Interaction: A Pharmacoepidemiologic Study. Clin Pharmacol Ther 102:547–553, 2017

49. Shah HR, Ledbetter L, Diasio R, et al: A retrospective study of coagulation abnormalities in patients receiving concomitant capecitabine and warfarin. Clin Colorectal Cancer 5:354–8, 2006

50. Malki MA, Pearson ER: Drug-drug-gene interactions and adverse drug reactions. Pharmacogenomics J 20:355–366, 2020

