## Supplement for "CLINICAL OPPORTUNITIES FOR GERMLINE PHARMACOGENETICS AND MANAGEMENT OF DRUG-DRUG INTERACTIONS IN PATIENTS WITH ADVANCED SOLID CANCERS"

**Supplemental Information**

**Supplemental Methods**

**Blood Sample Processing and DNA Genotyping**

Genomic DNA was extracted from subject whole blood sample using the EZ1^®^ kit from Qiagen (Germantown, MD) per the manufacturer's instructions. DNA samples were run on the QuantStudio™ 12K Flex (software v1.2.2; Waltham MA) for analysis. Taqman® allele discrimination was used to make genotype calls using either validated reagents or in a custom-designed methods using the OpenArray® platform (Thermo Fisher Scientific, Waltham, MA). In this method, genomic DNA was amplified and mixed with dual-labeled oligonucleotides that hybridize to the target sequence. The fluorescent reporter signal is released via hydrolysis by the 5’-3’ exonuclease activity of Taq polymerase, and quantitative measurement of the PCR product (via assessment of the fluorophore signal) is used to make genotype calls. The software that was used in these analyses includes Genotyper (v1.3) (Thermo Fisher Scientific, Waltham, MA) and Alleletyper™ (1.0) (Thermo Fisher Scientific, Waltham, MA)

*CYP2D6* copy number calling was performed using a TaqMan™ real-time polymerase chain reaction (PCR) targeting exon 9 (assay ID Hs00010001_cn) on a QuantStudio™ 12K Flex Real-Time PCR System (Thermo Fisher Scientific, Waltham, MA). The assay was performed within 96-well plates in four replicates along with an RNase P (assay ID 4403326, Thermo Fisher Scientific, Waltham, MA) reference control gene in accordance with the manufacturer’s instructions. To quantitate copy number, a two gene copy calibrator, a sample with one gene copy, and a sample with three gene copies were included in each run. CopyCaller™ Software v2.0 (Thermo Fisher Scientific, Waltham, MA) was used for relative quantification of *CYP2D6* copy number. If the calculated copy number for a probe was N ± 0.4 gene copies (where N is a whole number), it was predicted to be that whole number; conversely, if the calculated copy number was greater than ±0.4 gene copies, the result was assigned as a “no call,” and the assay was repeated. A confidence level of 95% and z-score value of <1.75 was applied to call the copy number.

**supplemental Tables**

**Table S1. List of all variants included on genotyping platform with minor allele frequencies in relevant ancestral populations.**

| **Gene** | **Allele** | **Variant** | **Function** | **Minor Allele Frequencies** |
| --- | --- | --- | --- | --- |
| ***CYP2B6*** | *6 | rs3745274 | Decreased | 0.32 (AA); 0.23 (E) |
|  | *18 | rs28399499 | None | 0.03 (AA); 0.00 (E) |
| ***CYP2C19*** | *2 | rs4244285 | None | 0.18 (AA); 0.12 (Am); 0.15 (E) |
|  | *3 | rs4986893 | None | 0.00 (AA); 0.00 (Am); 0.00 (E) |
|  | *4 | rs28399504 | None | 0.00 (AA); 0.00 (Am); 0.00 (E) |
|  | *6 | rs72552267 | None | 0.00 (AA); 0.00 (Am); 0.00 (E) |
|  | *8 | rs41291556 | None | 0.00 (AA); 0.00 (Am); 0.00 (E) |
|  | *17 | rs12248560 | Increased | 0.21 (AA); 0.09 (Am); 0.22 (E) |
| ***CYP2C9*** | *2 | rs1799853 | Decreased | 0.02 (AA); 0.03 (Am); 0.13 (E) |
|  | *3 | rs1057910 | None | 0.01 (AA); 0.03 (Am); 0.08 (E) |
|  | *5 | rs28371686 | Decreased | 0.01 (AA); 0.00 (Am); 0.00 (E) |
|  | *6 | rs9332131 | None | 0.01 (AA); 0.00 (Am); 0.00 (E) |
|  | *8 | rs7900194 | Decreased | 0.06 (AA); 0.02 (Am); 0.00 (E) |
|  | *11 | rs28371685 | Decreased | 0.01 (AA); 0.00 (Am); 0.00 (E) |
| ***CYP2D6*** | *2 | rs16947; rs1135840 | Normal | 0.16 (AA); 0.22 (Am); 0.28 (E) |
|  | *3 | rs35742686 | None | 0.00 (AA); 0.00 (Am); 0.02 (E) |
|  | *4 | rs3892097 | None | 0.05 (AA); 0.10 (Am); 0.19 (E) |
|  | *5 | Gene deletion | None | 0.05 (AA); 0.02 (Am); 0.03 (E) |
|  | *6 | rs5030655 | None | 0.00 (AA); 0.00 (Am); 0.01 (E) |
|  | *7 | rs5030867 | None | 0.00 (AA); 0.01 (Am); 0.00 (E) |
|  | *9 | rs5030656 | Decreased | 0.00 (AA); 0.00 (Am); 0.03 (E) |
|  | *10 | rs1065852; rs1135840 | Decreased | 0.04 (AA); 0.01 (Am); 0.02 (E) |
|  | *17 | rs28371706 | Decreased | 0.17 (AA); 0.00 (Am); 0.00 (E) |
|  | *29 | rs59421388 | Decreased | 0.09 (AA); 0.00 (Am); 0.00 (E) |
|  | *41 | rs28371725 | Decreased | 0.04 (AA); 0.02 (Am); 0.09 (E) |
| ***CYP3A4*** | *2 | rs55785340 | Decreased | 0.00 (AA); 0.00 (E) |
|  | *22 | rs35599367 | Decreased | 0.00 (AA); 0.05 (E) |
| ***CYP3A5*** | *3 | rs776746 | None | 0.32 (AA); 0.92 (E) |
|  | *6 | rs10264272 | None | 0.11 (AA); 0.00 (E) |
|  | *7 | rs41303343 | None | 0.12 (AA); 0.00 (E) |
| ***CYP4F2*** | *3 | rs2108622 | Decreased | 0.08 (AA); 0.41 (Am); 0.30 (E) |
| ***DPYD*** | *2A | rs3918290 | None | 0.00 (AA); 0.01 (E) |
|  | *9 | rs1801265 | Normal | 0.43 (AA); 0.23 (E) |
| ***G6PD*** | A | rs1050829 | Normal | 0.27 (AA); 0.00 (E) |
|  | A- | rs1050828 | Deficient | 0.11 (AA); 0.00 (E) |
| ***IFNL3*** |  | rs12979860 | Unfavorable | 0.60 (AA); 0.31 (E) |
| ***SLCO1B1*** | *5 | rs4149056 | Decreased | 0.00 (AA); 0.02 (E) |
|  | *17 | rs4149056; rs4149015 | Decreased | 0.05 (E) |
|  | *21 | rs4149015 | Decreased | 0.02 (E) |
| ***TPMT*** | *3A,*3B,*3C | rs1800460; rs1142345 | None | 0.02 (AA); 0.03 (E) |
| ***VKORC1*** |  | rs9923231 | Decreased | 0.10 (AA); 0.41 (E) |

**Abbreviations**: **AA** = African American; **Am** = American admixed; **E** = European

Allele frequencies are from PharmGKB gene-specific information tables (available at: https://www.pharmgkb.org/page/pgxGeneRef) or from the Allele Frequency Aggregator (ALFA) project as compiled in dbSNP (available at: https://www.ncbi.nlm.nih.gov/snp/).

**Table S2. List of all drugs included in the PGx analysis along with the assessed pharmacogenes and a list of all drugs included in the DDI analyses along with the affected CYP enzymes.**

| **Drugs Included in PGx Analyses** | | |
| --- | --- | --- |
| **Drug** | | **Assessed Pharmacogenes** |
| Amitriptyline | | *CYP2C19,* *CYP2D6* |
| Atomoxetine | | *CYP2D6* |
| Azathioprine | | *TPMT* |
| Capecitabine | | *DPYD* |
| Celecoxib | | *CYP2C9* |
| Citalopram | | *CYP2C19* |
| Clomipramine | | *CYP2C19*, *CYP2D6* |
| Clopidogrel | | *CYP2C19* |
| Codeine | | *CYP2D6* |
| Desipramine | | *CYP2D6* |
| Dexlansoprazole | | *CYP2C19* |
| Doxepin | | *CYP2C19, CYP2D6* |
| Efavirenz | | *CYP2B6* |
| Escitalopram | | *CYP2C19* |
| Fluorouracil | | *DPYD* |
| Flurbiprofen | | *CYP2C9* |
| Fluvoxamine | | *CYP2D6* |
| Ibuprofen | | *CYP2C9* |
| Imipramine | | *CYP2C19*, *CYP2D6* |
| Lansoprazole | | *CYP2C19* |
| Lornoxicam | | *CYP2C9* |
| Meloxicam | | *CYP2C9* |
| Mercaptopurine | | *TPMT* |
| Nortriptyline | | *CYP2D6* |
| Omeprazole | | *CYP2C19* |
| Ondansetron | | *CYP2D6* |
| Pantoprazole | | *CYP2C19* |
| Paroxetine | | *CYP2D6* |
| Peginterferon alfa-2a | | *IFNL3* |
| Peginterferon alfa-2b | | *IFNL3* |
| Phenytoin | | *CYP2C9* |
| Piroxicam | | *CYP2C9* |
| Rasburicase | | *G6PD* |
| Ribavirin | | *IFNL3* |
| Sertraline | | *CYP2C19* |
| Simvastatin | | *SLCO1B1* |
| Tacrolimus | | *CYP3A5* |
| Tamoxifen | | *CYP2D6* |
| Tegafur | | *DPYD* |
| Tenoxicam | | *CYP2C9* |
| Thioguanine | | *TPMT* |
| Tramadol | | *CYP2D6* |
| Trimipramine | | *CYP2C19, CYP2D6* |
| Tropisetron | | *CYP2D6* |
| Voriconazole | | *CYP2C19* |
| Warfarin | | *CYP2C9*, *CYP4F2*, *VKORC1* |
| **Drugs Included in CYP DDI Analyses** | | |
| **Drug** | **Association with Relevant CYP Enzymes** | |
| **Alfentanil** | **CYP3A Substrate** | |
| Allopurinol | CYP2C9 Inhibitor | |
| **Alprazolam** | **CYP3A Substrate** | |
| Amiodarone | CYP2C9 Inhibitor, CYP2D6 Inhibitor, CYP3A Inhibitor | |
| **Amitriptyline** | **CYP2C19 Substrate, CYP2D6 Substrate** | |
| Amlodipine | CYP3A Substrate | |
| Aprepitant | CYP3A Inhibitor | |
| **Aripiprazole** | **CYP2D6 Substrate** | |
| **Atomoxetine** | **CYP2D6 Substrate** | |
| Atorvastatin | CYP3A Substrate | |
| Boceprevir | CYP3A Inhibitor | |
| **Brexpiprazole** | **CYP2D6 Substrate** | |
| **Brivaracetam** | **CYP2C19 Substrate** | |
| **Bupropion** | **CYP2B6 Substrate**, CYP2D6 Inhibitor | |
| **Buspirone** | **CYP3A Substrate** | |
| Capecitabine | CYP2C9 Inhibitor | |
| **Carbamazepine** | **CYP3A Substrate,** CYP2B6 Inducer, CYP2C19 Inducer, CYP2C9 Inducer, CYP3A Inducer | |
| **Carisoprodol** | **CYP2C19 Substrate** | |
| Carvedilol | CYP2D6 Substrate | |
| Celecoxib | CYP2C9 Substrate, CYP2D6 Inhibitor | |
| Cerivastatin | CYP2C8 Substrate | |
| Cevimeline | CYP2D6 Substrate | |
| Chlorpheniramine | CYP2D6 Inhibitor | |
| Chlorpromazine | CYP2D6 Inhibitor | |
| Cimetidine | CYP2C9 Inhibitor, CYP2D6 Inhibitor | |
| Cinacalcet | CYP2D6 Inhibitor | |
| Ciprofloxacin | CYP3A Inhibitor | |
| **Citalopram** | **CYP2C19 Substrate** | |
| **Clarithromycin** | **CYP3A Substrate**, CYP3A Inhibitor | |
| **Clobazam** | **CYP2C19 Substrate** | |
| **Clomipramine** | **CYP2C19 Substrate, CYP2D6 Substrate** | |
| **Clopidogrel** | **CYP2C19 Substrate** | |
| **Clozapine** | **CYP2D6 Substrate** | |
| Cobicistat | CYP3A Inhibitor | |
| **Codeine** | **CYP2D6 Substrate** | |
| **Copanlisib** | **CYP3A Substrate** | |
| Dasabuvir | CYP2C8 Substrate, CYP3A Inhibitor | |
| **Desipramine** | **CYP2D6 Substrate** | |
| **Deutetrabenazine** | **CYP2D6 Substrate** | |
| Dexamethasone | CYP2C19 Inducer, CYP2C9 Inducer, CYP3A Inducer | |
| Dexlansoprazole | CYP2C19 Substrate | |
| Dextromethorphan | CYP2D6 Substrate | |
| Diltiazem | CYP3A Substrate, CYP3A Inhibitor | |
| Diphenhydramine | CYP2D6 Inhibitor | |
| **Docetaxel** | **CYP3A Substrate** | |
| **Donepezil** | **CYP2D6 Substrate** | |
| **Doxepin** | **CYP2C19 Substrate,** CYP2D6 Substrate, CYP2D6 Inhibitor | |
| Doxorubicin | CYP2D6 Inhibitor | |
| **Dronabinol** | **CYP2C9 Substrate** | |
| Duloxetine | CYP2D6 Inhibitor | |
| Efavirenz | CYP2B6 Substrate, CYP2B6 Inducer, CYP2C19 Inducer, CYP3A Inducer | |
| Eliglustat | CYP2D6 Substrate | |
| **Enzalutamide** | **CYP2C8 Substrate**, CYP2C19 Inducer, CYP2C9 Inducer, CYP3A Inducer | |
| **Erdafitinib** | **CYP2C9 Substrate** | |
| **Erythromycin** | **CYP3A Substrate,** CYP3A Inhibitor | |
| **Escitalopram** | **CYP2C19 Substrate** | |
| Esomeprazole | CYP2C19 Substrate, CYP2C19 Inhibitor | |
| Felbamate | CYP2C19 Inhibitor | |
| Fenofibrate | CYP2C9 Inhibitor | |
| **Fentanyl** | **CYP3A Substrate** | |
| Flibanserin | CYP2C19 Substrate | |
| Fluconazole | CYP2C19 Inhibitor, CYP2C9 Inhibitor, CYP3A Inhibitor | |
| Fluoxetine | CYP2C19 Inhibitor, CYP2D6 Inhibitor | |
| Flurbiprofen | CYP2C9 Substrate | |
| Fluvastatin | CYP2C9 Inhibitor | |
| **Fluvoxamine** | **CYP2D6 Substrate,** CYP2C19 Inhibitor, CYP2C9 Inhibitor | |
| **Gefitinib** | **CYP2D6 Substrate** | |
| Gemfibrozil | CYP2C8 Inhibitor | |
| Halofantrine | CYP2D6 Inhibitor | |
| **Haloperidol** | **CYP3A Substrate,** CYP2D6 Inhibitor | |
| Ibuprofen | CYP2C9 Substrate | |
| **Iloperidone** | **CYP2D6 Substrate** | |
| **Imatinib** | **CYP3A Substrate,** CYP3A Inhibitor | |
| **Imipramine** | **CYP2C19 Substrate, CYP2D6 Substrate** | |
| Indinavir | CYP3A Inhibitor | |
| Isoniazid | CYP2C19 Inhibitor | |
| **Ketamine** | **CYP2B6 Substrate** | |
| Ketoconazole | CYP3A Inhibitor | |
| Lansoprazole | CYP2C19 Substrate | |
| Lofexidine | CYP2D6 Substrate | |
| Lornoxicam | CYP2C9 Substrate | |
| Meclizine | CYP2D6 Substrate | |
| Meloxicam | CYP2C9 Substrate | |
| **Methadone** | **CYP2B6 Substrate,** CYP2D6 Inhibitor | |
| **Methylphenidate** | **CYP2D6 Substrate** | |
| Methylprednisolone | CYP2C19 Inducer, CYP2C9 Inducer, CYP3A Inducer | |
| Metoclopramide | CYP2D6 Substrate | |
| Metronidazole | CYP2C9 Inhibitor | |
| **Midazolam** | **CYP3A Substrate** | |
| Midodrine | CYP2D6 Inhibitor | |
| Mifepristone | CYP3A Inhibitor | |
| Mirabegron | CYP2D6 Inhibitor | |
| **Mirtazapine** | **CYP2D6 Substrate** | |
| Nefazodone | CYP3A Inhibitor | |
| Nelfinavir | CYP3A Inhibitor | |
| Nevirapine | CYP2B6 Substrate, CYP2C19 Inducer, CYP2C9 Inducer, CYP3A Inducer | |
| Nifedipine | CYP3A Substrate | |
| **Norfloxacin** | **CYP3A Inhibitor** | |
| **Nortriptyline** | **CYP2D6 Substrate** | |
| **Olaparib** | **CYP3A Substrate** | |
| Omeprazole | CYP2C19 Substrate, CYP2C19 Inhibitor | |
| Ondansetron | CYP2D6 Substrate | |
| Oxcarbazepine | CYP2C19 Inhibitor, CYP2C19 Inducer, CYP2C9 Inducer, CYP3A Inducer | |
| **Oxycodone** | **CYP2D6 Substrate** | |
| **Paclitaxel** | **CYP2C8 Substrate** | |
| **Palbociclib** | **CYP3A Substrate** | |
| Pantoprazole | CYP2C19 Substrate | |
| **Paroxetine** | **CYP2D6 Substrate,** CYP2D6 Inhibitor | |
| **Perphenazine** | **CYP2D6 Substrate,** CYP2D6 Inhibitor | |
| Phenobarbital | CYP2B6 Inducer, CYP2C19 Inducer, CYP2C9 Inducer, CYP3A Inducer | |
| **Phenytoin** | **CYP2C9 Substrate,** CYP2C19 Inducer, CYP2C9 Inducer, CYP3A Inducer | |
| **Pimozide** | **CYP2D6 Substrate** | |
| Pioglitazone | CYP2C8 Substrate | |
| Piroxicam | CYP2C9 Substrate | |
| **Pitolisant** | **CYP2D6 Substrate** | |
| Prednisone | CYP2C19 Inducer, CYP2C9 Inducer, CYP3A Inducer | |
| Promethazine | CYP2D6 Inhibitor | |
| **Propafenone** | **CYP2D6 Substrate** | |
| **Propofol** | **CYP2B6 Substrate** | |
| **Protriptyline** | **CYP2D6 Substrate** | |
| **Quinidine** | **CYP2D6 Substrate,** CYP2D6 Inhibitor | |
| Rabeprazole | CYP2C19 Substrate | |
| **Regorafenib** | **CYP3A Substrate** | |
| Repaglinide | CYP2C8 Substrate | |
| Rifabutin | CYP2C19 Inducer, CYP2C9 Inducer, CYP3A Inducer | |
| Rifampin | CYP2B6 Inducer, CYP2C8 Inducer, CYP2C19 Inducer, CYP2C9 Inducer, CYP3A Inducer | |
| **Risperidone** | **CYP2D6 Substrate** | |
| Ritonavir | CYP2D6 Inhibitor, CYP3A Inhibitor | |
| **Sertraline** | **CYP2C19 Substrate, CYP2D6 Substrate,** CYP2C9 Inhibitor, CYP2D6 Inhibitor | |
| Sildenafil | CYP3A Substrate | |
| Simvastatin | CYP3A Substrate | |
| Siponimod | CYP2C9 Substrate | |
| **Sirolimus** | **CYP3A Substrate** | |
| **Sorafenib** | **CYP3A Substrate** | |
| St. John's Wort | CYP2C19 Inducer, CYP2C9 Inducer, CYP3A Inducer | |
| **Sunitinib** | **CYP3A Substrate** | |
| Suvorexant | CYP3A Substrate | |
| **Tacrolimus** | **CYP3A Substrate** | |
| Tafenoquine | CYP2C9 Substrate | |
| **Tamoxifen** | **CYP2D6 Substrate** | |
| Telaprevir | CYP3A Inhibitor | |
| Teniposide | CYP2C9 Inhibitor | |
| Terbinafine | CYP2D6 Inhibitor | |
| Teriflunomide | CYP2C8 Inhibitor | |
| **Tetrabenazine** | **CYP2D6 Substrate** | |
| **Thioridazine** | **CYP2D6 Substrate,** CYP2D6 Inhibitor | |
| Thiotepa | CYP2B6 Inhibitor | |
| Tolterodine | CYP2D6 Substrate | |
| Topiramate | CYP2C19 Inhibitor | |
| Torsemide | CYP2C8 Substrate | |
| **Tramadol** | **CYP3A Substrate, CYP2D6 Substrate** | |
| **Trazodone** | **CYP3A Substrate** | |
| Trimethoprim | CYP2C9 Inhibitor | |
| **Trimipramine** | **CYP2C19 Substrate, CYP2D6 Substrate** | |
| **Valbenazine** | **CYP2D6 Substrate** | |
| **Vemurafenib** | **CYP3A Substrate** | |
| **Venlafaxine** | **CYP2D6 Substrate** | |
| Verapamil | CYP3A Substrate, CYP3A Inhibitor | |
| **Vincristine** | **CYP3A Substrate** | |
| **Voriconazole** | **CYP2C19 Substrate, CYP3A Substrate,** CYP2B6 Inhibitor, CYP2C19 Inhibitor, CYP2C9 Inhibitor, CYP3A Inhibitor | |
| **Vortioxetine** | **CYP2D6 Substrate** | |
| **Warfarin** | **CYP2C9 Substrate** | |
| **Drugs Included in Tyrosine Kinase Inhibitor-Acid Reducer DDI Analyses** | | |
| Acid Reducers | Aluminum hydroxide; calcium carbonate; cimetidine; dexlansoprazole; esomeprazole; famotidine; lansoprazole; magnesium carbonate; magnesium hydroxide; nizatidine; omeprazole; pantoprazole; rabeprazole; ranitidine; sodium bicarbonate; sucralfate | |
| Tyrosine Kinase Inhibitors | Acalabrutinib; afatinib; alectinib; avapritinib; axitinib; bosutinib; cabozantinib; crizotinib; dacomitinib; dasatinib; entrectinib; erlotinib; gilteritinib; ibrutinib; imatinib; lapatinib; midostaurin; neratinib; nilotinib; pacritinib; pazopanib; pexidartinib; ponatinib; regorafenib; sorafenib; sunitinib; tucatinib; vandetanib; zanubritinib; ziv-aflibercept | |

Bolded drugs indicate that these medications were classified as “serious DDIs” when they were involved as sensitive substrates in DDIs.

**Table S3. Days supply assumed for prescriptions dispensed from a pharmacy (used to determine concomitant administration of CYP inducer, inhibitor, and substrates drugs within DDI analyses).**

| **Drug** | **Assumed Days Supply** |
| --- | --- |
| Alfentanil | 1 |
| Allopurinol | 30 |
| Alprazolam | 7 |
| Amiodarone | 7 |
| Amitriptyline | 30 |
| Amlodipine | 30 |
| Aprepitant | 4 |
| Aripiprazole | 30 |
| Atomoxetine | 30 |
| Atorvastatin | 30 |
| Boceprevir | 30 |
| Brexpiprazole | 30 |
| Brivaracetam | 30 |
| Bupropion | 30 |
| Buspirone | 30 |
| Capecitabine | 14 |
| Carbamazepine | 30 |
| Carisoprodol | 7 |
| Carvedilol | 30 |
| Celecoxib | 7 |
| Cerivastatin | 30 |
| Cevimeline | 30 |
| Chlorpheniramine | 1 |
| Chlorpromazine | 1 |
| Cimetidine | 14 |
| Cinacalcet | 30 |
| Ciprofloxacin | 7 |
| Citalopram | 30 |
| Clarithromycin | 7 |
| Clobazam | 30 |
| Clomipramine | 30 |
| Clopidogrel | 30 |
| Clozapine | 30 |
| Cobicistat | 30 |
| Codeine | 7 |
| Copanlisib | 28 |
| Dasabuvir | 30 |
| Desipramine | 30 |
| Deutetrabenazine | 30 |
| Dexamethasone | 7 |
| Dexlansoprazole | 14 |
| Dextromethorphan | 1 |
| Diltiazem | 30 |
| Diphenhydramine | 1 |
| Docetaxel | 1 |
| Donepezil | 30 |
| Doxepin | 30 |
| Doxorubicin | 1 |
| Dronabinol | 1 |
| Duloxetine | 30 |
| Efavirenz | 30 |
| Eliglustat | 30 |
| Enzalutamide | 30 |
| Erdafitinib | 30 |
| Erythromycin | 7 |
| Escitalopram | 30 |
| Esomeprazole | 14 |
| Felbamate | 30 |
| Fenofibrate | 30 |
| Fentanyl | 7 |
| Flibanserin | 30 |
| Fluconazole | 7 |
| Fluoxetine | 30 |
| Flurbiprofen | 7 |
| Fluvastatin | 30 |
| Fluvoxamine | 30 |
| Gefitinib | 30 |
| Gemfibrozil | 30 |
| Halofantrine | 7 |
| Haloperidol | 1 |
| Ibuprofen | 7 |
| Iloperidone | 30 |
| Imatinib | 30 |
| Imipramine | 30 |
| Indinavir | 30 |
| Isoniazid | 30 |
| Ketamine | 1 |
| Ketoconazole | 7 |
| Lansoprazole | 14 |
| Lofexidine | 7 |
| Lornoxicam | 7 |
| Meclizine | 1 |
| Meloxicam | 7 |
| Methadone | 7 |
| Methylphenidate | 30 |
| Methylprednisolone | 7 |
| Metoclopramide | 14 |
| Metronidazole | 7 |
| Midazolam | 1 |
| Midodrine | 30 |
| Mifepristone | 30 |
| Mirabegron | 30 |
| Mirtazapine | 30 |
| Nefazodone | 30 |
| Nelfinavir | 30 |
| Nevirapine | 30 |
| Nifedipine | 30 |
| Norfloxacin | 7 |
| Nortriptyline | 30 |
| Olaparib | 30 |
| Omeprazole | 14 |
| Ondansetron | 2 |
| Oxcarbazepine | 30 |
| Oxycodone | 7 |
| Paclitaxel | 1 |
| Palbociclib | 21 |
| Pantoprazole | 14 |
| Paroxetine | 30 |
| Perphenazine | 30 |
| Phenobarbital | 30 |
| Phenytoin | 30 |
| Pimozide | 30 |
| Pioglitazone | 30 |
| Piroxicam | 7 |
| Pitolisant | 30 |
| Prednisone | 7 |
| Promethazine | 1 |
| Propafenone | 1 |
| Propofol | 1 |
| Protriptyline | 30 |
| Quinidine | 1 |
| Rabeprazole | 14 |
| Regorafenib | 21 |
| Repaglinide | 30 |
| Rifabutin | 30 |
| Rifampin | 30 |
| Risperidone | 30 |
| Ritonavir | 30 |
| Sertraline | 30 |
| Sildenafil | 1 |
| Simvastatin | 30 |
| Siponimod | 30 |
| Sirolimus | 30 |
| Sorafenib | 30 |
| St. John's Wort | 30 |
| Sunitinib | 28 |
| Suvorexant | 30 |
| Tacrolimus | 30 |
| Tafenoquine | 17 |
| Tamoxifen | 30 |
| Telaprevir | 30 |
| Teniposide | 1 |
| Terbinafine | 30 |
| Teriflunomide | 30 |
| Tetrabenazine | 30 |
| Thioridazine | 30 |
| Thiotepa | 1 |
| Tolterodine | 30 |
| Topiramate | 30 |
| Torsemide | 30 |
| Tramadol | 7 |
| Trazodone | 30 |
| Trimethoprim | 7 |
| Trimipramine | 30 |
| Valbenazine | 30 |
| Vemurafenib | 30 |
| Venlafaxine | 30 |
| Verapamil | 30 |
| Vincristine | 1 |
| Voriconazole | 7 |
| Vortioxetine | 30 |
| Warfarin | 30 |

**Table S4. Most common drug-drug pairs contained within DDIs by enzyme involved, including (left) and excluding (right) DDIs involving corticosteroids.**

| **Enzyme** | **DDIs Including Corticosteroids** | | **DDIs Excluding Corticosteroids** | |
| --- | --- | --- | --- | --- |
|  | **Drug-Drug Pair (Perpetrator-Victim)** | **Frequency** | **Drug-Drug Pair (Perpetrator-Victim)** | **Frequency** |
| CYP2B6 | **Carbamazepine-Methadone** | 1 | **Carbamazepine-Methadone** | 1 |
| CYP2C19 | Dexamethasone-Pantoprazole | 33 | Fluconazole-Pantoprazole | 12 |
|  | Dexamethasone-Omeprazole | 18 | **Omeprazole-Sertraline** | 9 |
|  | **Dexamethasone-Escitalopram** | 15 | Fluconazole-Omeprazole | 7 |
|  | **Dexamethasone-Sertraline** | 13 | **Fluconazole-Escitalopram; Fluconazole-Sertraline;**  Fluoxetine-Pantoprazole* | 4 |
|  | Fluconazole-Pantoprazole | 12 |  |  |
| CYP2C9 | Dexamethasone-Ibuprofen | 10 | Metronidazole-Ibuprofen | 6 |
|  | **Dexamethasone-Warfarin** | 9 | Sertraline-Ibuprofen | 4 |
|  | Metronidazole-Ibuprofen | 6 | **Sertraline-Dronabinol** | 3 |
|  | **Dexamethasone-Dronabinol;**  **Prednisone-Warfarin;**  Sertraline-Ibuprofen | 4 | Allopurinol-Ibuprofen;  **Allopurinol-Meloxicam;**  Fluconazole-Ibuprofen;  **Fluconazole-Warfarin^+^** | 2 |
|  | **Sertraline-Dronabinol** | 3 |  |  |
| CYP2D6 | Promethazine-Ondansetron | 174 | Promethazine-Ondansetron | 174 |
|  | Diphenhydramine-Ondansetron | 115 | Diphenhydramine-Ondansetron | 115 |
|  | Haloperidol-Ondansetron | 46 | Haloperidol-Ondansetron | 46 |
|  | Promethazine-Metoclopramide | 35 | Promethazine-Metoclopramide | 35 |
|  | Diphenhydramine-Metoclopramide | 19 | Diphenhydramine-Metoclopramide | 19 |
| CYP3A | **Ciprofloxacin-Fentanyl** | 40 | **Ciprofloxacin-Fentanyl** | 40 |
|  | **Ciprofloxacin-Midazolam** | 25 | **Ciprofloxacin-Midazolam** | 25 |
|  | **Dexamethasone-Fentanyl** | 23 | **Fluconazole-Fentanyl** | 13 |
|  | Dexamethasone-Atorvastatin | 17 | **Ciprofloxacin-Tramadol;**  **Diltiazem-Fentanyl** | 9 |
|  | Dexamethasone-Amlodipine | 14 | Ciprofloxacin-Amlodipine;  **Fluconazole-Midazolam** | 7 |

Note: Bolded DDI pairs contained substrate drugs with narrow therapeutic indices or the potential for serious adverse drug events and were included as “serious DDIs.”

* When corticosteroids were excluded, additional CYP2C19 DDI pairs with a frequency of 4 included: omeprazole-escitalopram and topiramate-pantoprazole.

^+^When corticosteroids were excluded, additional CYP2C9 DDI pairs with a frequency of 2 included: metronidazole-meloxicam and metronidazole-warfarin.

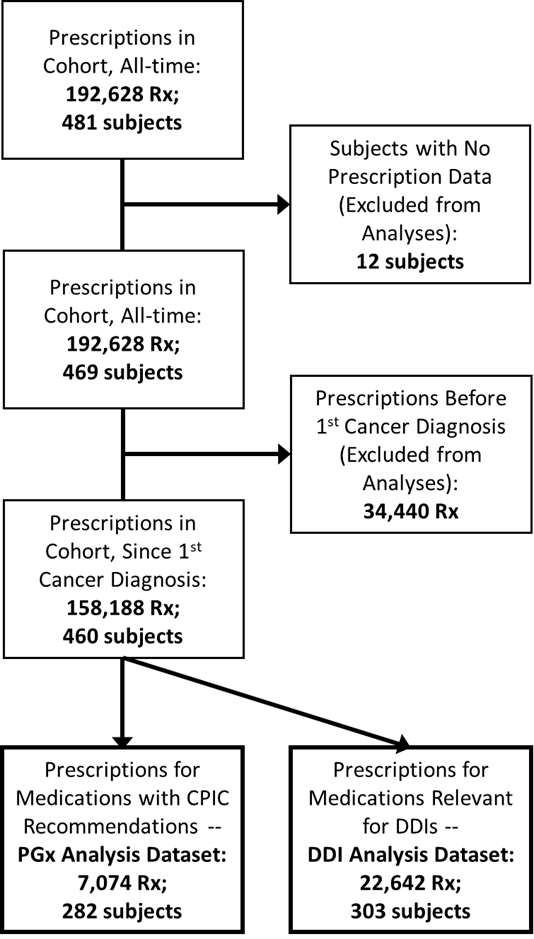
**supplemental figures**

**Figure S1. Diagram of medication filtering workflow, including unique medications and unique subjects included in the PGx and DDI analyses.**

**Abbreviations**: **Rx** = unit to denote number of prescriptions

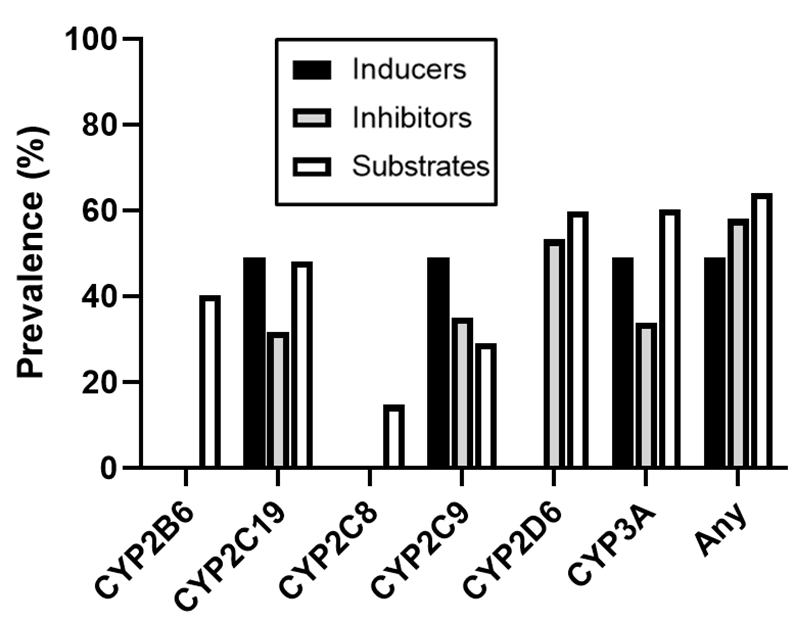
**Figure S2. Subject-level prevalence of prescriptions for inducers, inhibitors, and inducers of major CYP enzymes.**
